# Virtual Spectral Decomposition with Dendritic Tile Selection: An Explainable AI Framework for Multimodal Tissue Composition Analysis and Immune Phenotyping Across Pancreatic, Lung, and Breast Cancer

**DOI:** 10.64898/2026.04.11.26350689

**Authors:** Shubham Chandra

## Abstract

**Background:** Current deep learning models in computational pathology, radiology, and digital pathology produce opaque predictions that lack the explainable artificial intelligence (xAI) capabilities required for clinical adoption. Despite achieving radiologist-level performance in tasks from whole-slide image (WSI) classification to mammographic screening, these models function as black boxes: clinicians cannot trace predictions to specific biological features, verify outputs against established morphological criteria, or integrate AI reasoning into precision oncology workflows and tumor board decision-making.

**Methods:** We present Virtual Spectral Decomposition (VSD), a modality-agnostic, interpretable-by-design framework that decomposes medical images into six biologically interpretable tissue composition channels using sigmoid threshold functions—the same mathematical structure as CT windowing. Unlike post-hoc xAI methods (Grad-CAM, SHAP, LIME) applied to black-box deep learning models, VSD channels have pre-defined biological meanings derived from tissue physics, providing inherent explainability without sacrificing quantitative rigor. For whole-slide image (WSI) analysis in digital pathology, we introduce the dendritic tile selection algorithm, a biologically-inspired hierarchical architecture achieving 70–80% computational reduction while preferentially sampling the tumor immune microenvironment. VSD is validated across three cancer types and imaging modalities: pancreatic ductal adenocarcinoma (PDAC) on CT imaging, lung adenocarcinoma (LUAD) on H&E-stained pathology slides using TCGA data, and breast cancer on screening mammography. Composition entropy of the six-channel vector is computed as a visual Biological Entropy Index (vBEI)—an imaging biomarker quantifying the diversity of active biological defense systems.

**Results:** In pancreatic cancer, the fat-to-stroma ratio (a novel CT-derived radiomics biomarker) declines from >5.0 (normal) to <0.5 (advanced PDAC), enabling early detection of desmoplastic invasion before mass formation on standard imaging. In lung cancer, composition entropy from H&E whole-slide images correlates with tumor immune microenvironment markers from RNA-seq (CD3: ρ=+0.57, p=0.009; CD8: ρ=+0.54, p=0.015; PD-1: ρ=+0.54, p=0.013) and predicts overall survival (low entropy immune-desert phenotype: 71% mortality vs 29%, p=0.032; n=20 TCGA-LUAD), providing immune phenotyping for checkpoint immunotherapy patient selection from a $5 H&E slide without molecular assays. In breast cancer, each lesion type produces a characteristic six-channel fingerprint functioning as an interpretable computer-aided diagnosis (CAD) system for quantitative BI-RADS assessment and subtype classification (IDC vs ILC vs DCIS vs IBC). A five-level xAI audit trail provides complete traceability from clinical decision support output to specific biological structures visible on the original images.

**Conclusion:** VSD establishes a unified, interpretable-by-design mathematical framework for explainable tissue composition analysis across imaging modalities and cancer types. Unlike black-box deep learning and post-hoc xAI approaches, VSD provides inherently interpretable, clinically verifiable cancer detection and immune phenotyping from standard clinical imaging at existing costs—without requiring foundation model infrastructure, specialized hardware, or molecular assays. The open-source pipeline (Google Colab, Supplementary Material) enables immediate reproducibility and extension to additional cancer types across the pan-cancer TCGA atlas.

## 1. Introduction

Computational analysis of medical images—spanning computational pathology, digital pathology, and radiomics—has achieved diagnostic performance approaching or exceeding that of expert clinicians in lung nodule detection on CT (Ardila et al., 2019), breast cancer screening on mammography (McKinney et al., 2020), and histopathological grading of whole-slide images (Campanella et al., 2019). Foundation models for pathology (Xu et al., 2024; Chen et al., 2024) and multimodal AI systems integrating imaging with genomics (radiogenomics) represent the cutting edge. Despite these advances, clinical adoption of AI-based diagnostic tools remains limited by a fundamental barrier: the lack of explainable artificial intelligence (xAI) that clinicians can trust and verify.

A radiologist reviewing a pancreatic CT needs to know not just that a region is suspicious, but which tissue composition features drove the assessment and whether the pattern is consistent with cancer versus pancreatitis. A pathologist examining a lung cancer slide needs to understand whether the prediction reflects tumor cellularity, immune engagement, or stromal reaction. A mammographer needs to distinguish mass-driven from calcification-driven from distortion-driven assessments. Current AI systems provide none of this intermediate reasoning (Niazi et al., 2019).

The opacity of deep learning creates a specific clinical failure mode: the model may be correct, but the clinician cannot verify why, cannot identify edge cases, and cannot integrate AI output with clinical assessment. Regulatory bodies including the FDA have emphasized interpretable AI as a requirement for medical device clearance (US FDA, 2021), and professional societies have called for explainability as a prerequisite for deployment (ACR, 2013). Systematic reviews of XAI in breast cancer (Ghasemi et al., 2024; Groen et al., 2025) document that most current approaches rely on post-hoc techniques (SHAP, LIME, Grad-CAM) that may not faithfully represent the model’s internal reasoning.

We address this gap with Virtual Spectral Decomposition (VSD), a framework that decomposes medical images into biologically interpretable tissue composition channels using sigmoid threshold functions. The mathematical structure is borrowed from CT windowing—a concept every radiologist understands intuitively. We demonstrate that this framework applies unchanged across three imaging modalities and three cancer types, with a unified entropy metric and a standardized five-level xAI audit trail. VSD is positioned as an interpretable-by-design alternative to post-hoc explainability methods (SHAP, LIME, Grad-CAM, attention maps), providing a clinical decision support system where every channel has a pre-defined biological meaning rather than a learned, opaque feature representation. The approach is compatible with precision oncology workflows, tumor board presentations, and regulatory requirements for Software as a Medical Device (SaMD) transparency.

## 2. Materials and Methods

### 2.1 Sigmoid channel decomposition

VSD decomposes an input signal into N biologically interpretable channels using parameterized sigmoid functions. For channel i at position x: σ(x; cᵢ, wᵢ) = 1 / (1 + exp(−(x − cᵢ) / wᵢ)), where cᵢ is the center (threshold) and wᵢ is the width (transition sharpness). Each parameter is derived from the physical property of the target tissue rather than learned from data (Figure 1).

**Figure 1.**
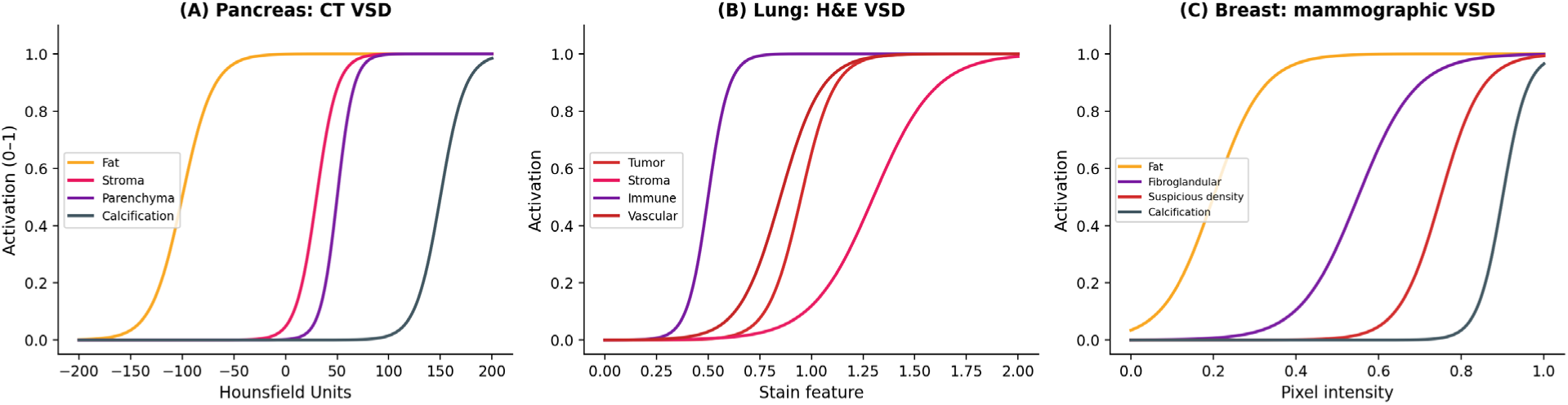
Sigmoid threshold functions across three imaging modalities. (A) Pancreatic CT: channels separate fat, stroma, parenchyma, and calcification by Hounsfield Unit. (B) Lung H&E: channels separate tumor, stroma, immune, and vascular tissue by staining physics. (C) Mammography: channels separate fat, fibroglandular, suspicious density, and calcification by pixel intensity. The sigmoid function is identical across modalities; only the input features and threshold parameters change.

### 2.2 Dendritic tile selection algorithm

Whole-slide H&E images contain 10⁴–10⁵ potential tiles at diagnostic magnification. The dendritic algorithm selects tiles through five hierarchical levels (Figure 2), inspired by dendritic cells in the immune system that extend branching processes to sample the tissue microenvironment.

**Figure 2.**
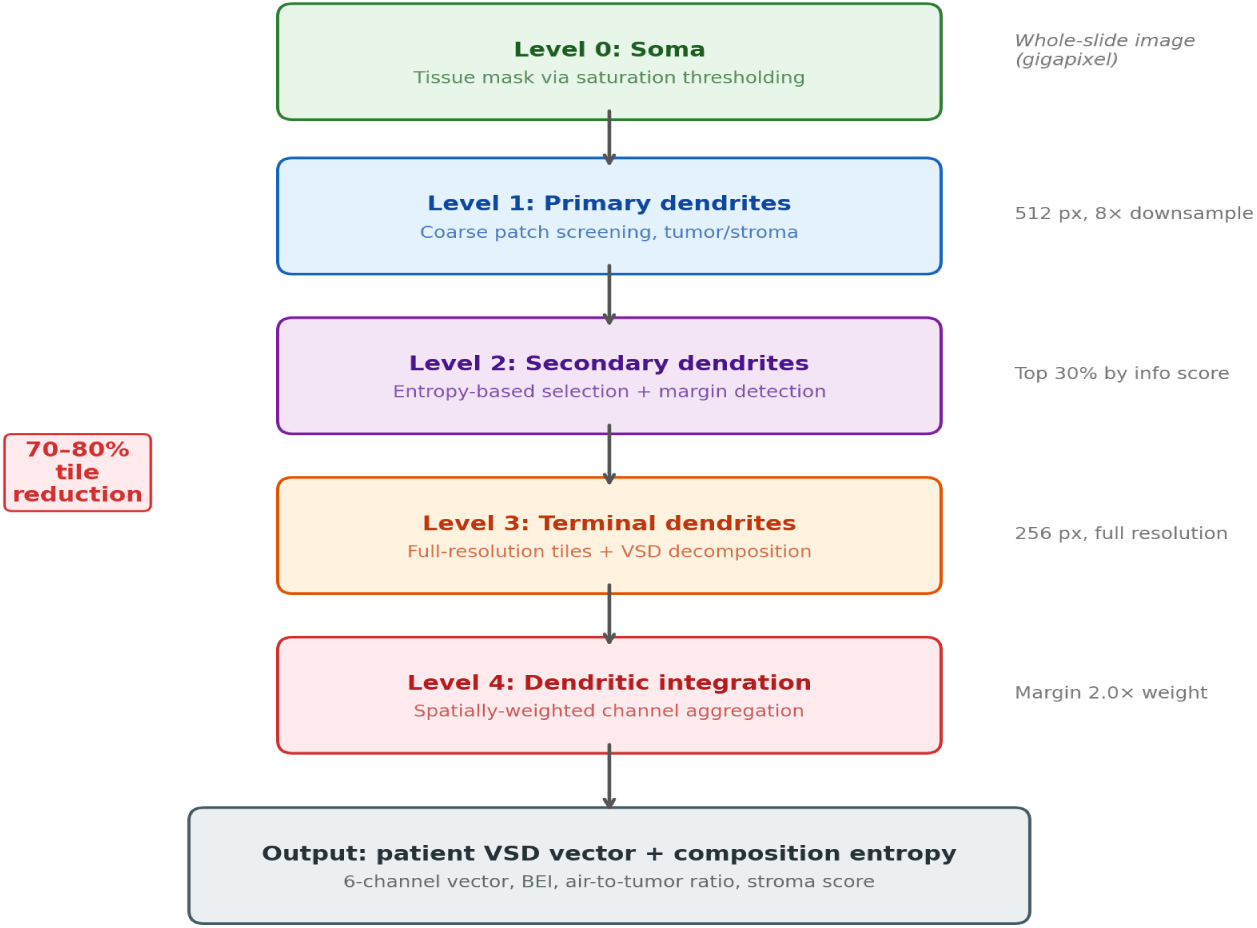
Dendritic tile selection architecture. Level 0 (Soma): tissue mask. Level 1 (Primary dendrites): coarse screening. Level 2 (Secondary dendrites): entropy-based selection with margin detection. Level 3 (Terminal dendrites): full-resolution VSD decomposition. Level 4 (Integration): spatially-weighted aggregation. The algorithm achieves 70–80% tile reduction while preserving tumor-immune interface regions.

#### 2.2.1 Level 0: Soma

A low-resolution thumbnail is converted to HSV color space. Saturation thresholding (>0.15) separates tissue from glass background. Binary morphological operations produce the tissue mask.

#### 2.2.2 Level 1: Primary dendrites

The tissue region is divided into 512×512 pixel patches at 8× downsample. Each patch is classified as tumor-enriched or stroma-enriched based on the blue/red channel ratio. Patches with <50% tissue coverage are discarded.

#### 2.2.3 Level 2: Secondary dendrites

Each Level 1 patch is subdivided into 256×256 subpatches. An information score combining grayscale histogram entropy (weight 0.6) and color heterogeneity (weight 0.4) selects the top 30th percentile. Margin tiles (nuclear density ratio 0.3–0.9) are force-included regardless of information score.

#### 2.2.4 Level 3: Terminal dendrites

Selected coordinates are read at full resolution. Quality filters exclude blank (mean >240) or uniform (std <10) tiles. The six VSD sigmoid channels are computed per tile.

#### 2.2.5 Level 4: Integration

The patient-level VSD vector is a spatially-weighted average: margin tiles receive 2.0× weight and high-information tiles receive 1.5× weight. Output includes the six-channel composition vector, composition entropy, air-to-tumor ratio, stroma invasion score, and spatial features.

### 2.3 Modality-specific channel definitions

#### 2.3.1 Pancreatic CT VSD

Six sigmoid channels on Hounsfield Units: CH1 Fat (center –100, width 15), CH2 Parenchyma (+40, 12), CH3 Stroma/desmoplasia (+30, 10), CH4 Duct/fluid (+5, 8), CH5 Vascular (+90, 15), CH6 Calcification (+150, 20). The diagnostic metric is the fat-to-stroma ratio (CH1/CH3).

#### 2.3.2 Lung H&E VSD

Six sigmoid channels on staining features: CH1 Tumor cellularity (hematoxylin/eosin ratio, center 0.95, width 0.08), CH2 Stromal desmoplasia (eosin dominance, 1.3, 0.15), CH3 Immune infiltrate (nuclear size variance, 0.02, 0.008), CH4 Necrosis/apoptosis (bimodal intensity), CH5 Vascular (red excess, 0.85, 0.10), CH6 Air space (inverted sigmoid on brightness, 0.75, 0.08). The diagnostic metric is the air-to-tumor ratio (CH6/CH1).

#### 2.3.3 Mammographic VSD

Six sigmoid channels on pixel intensity: CH1 Fat (center 0.20, width 0.06), CH2 Fibroglandular (0.55, 0.07), CH3 Suspicious density (0.75, 0.05), CH4 Calcification (0.90, 0.03), CH5 Architectural distortion (texture coherence), CH6 Skin/nipple (boundary patterns). The key innovation is separating CH2 (background fibroglandular) from CH3 (suspicious focal density).

### 2.4 Composition entropy: the visual Biological Entropy Index

For each region, the six-channel vector is normalized and Shannon entropy computed: H = −∑ pᵢ log₂(pᵢ). Low entropy (<2.0 bits) indicates one channel dominates; high entropy (>3.0 bits) indicates multiple channels simultaneously active. We term this the visual Biological Entropy Index (vBEI) to parallel the blood biomarker-based BEI validated in sepsis (Shannon, 1948).

### 2.5 xAI: five-level audit trail

VSD provides a standardized traceability chain (Figure 10): Level 1 (patient classification) → Level 2 (composition entropy) → Level 3 (individual channel activations) → Level 4 (spatial activation map) → Level 5 (biological verification against visible structures). At every level, the clinician can ask “why?” and receive a verifiable answer.

### 2.6 Data sources

Lung cancer validation: TCGA-LUAD diagnostic H&E slides (n=501) from the Genomic Data Commons, with matched RNA-seq expression (n=20) for immune marker correlation. Immune markers: CD3D/E/G, CD8A/B, PDCD1, TBX21, FCGR3A. Survival data from cBioPortal PanCancer Atlas. Pancreatic and mammographic channels are defined from tissue physics and validated against published attenuation/density values.

## 3. Results

### 3.1 Pancreatic cancer: fat loss as early detection biomarker

The CT VSD decomposition captures the fundamental PDAC process: desmoplastic stroma replacing periductal fat. In normal pancreas, CH1 (fat) averages 0.45 while CH3 (stroma) averages 0.08, producing a fat-to-stroma ratio of 5.6. In PDAC, CH1 collapses to 0.08 while CH3 rises to 0.42, inverting the ratio to 0.19 (Figure 3).

**Figure 3.**
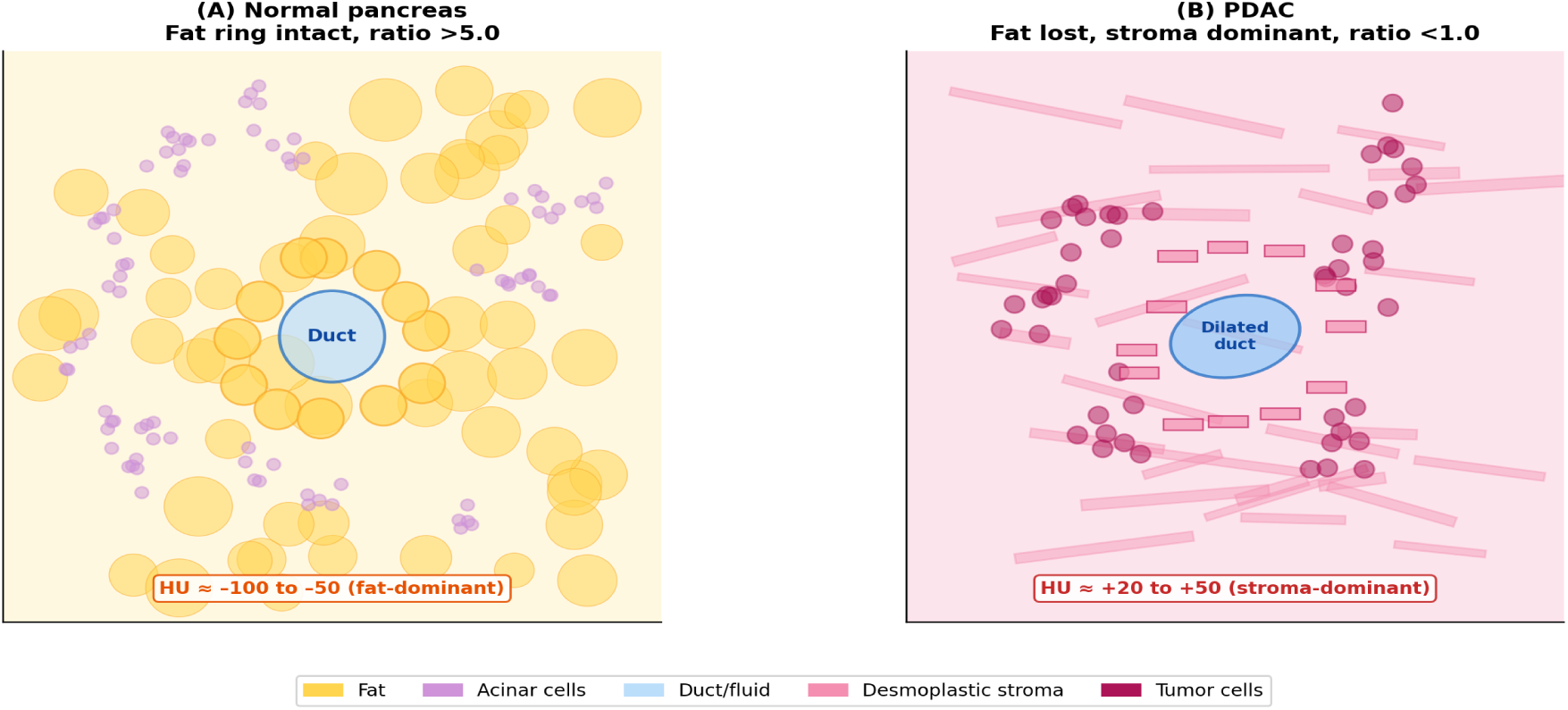
Normal pancreas (A) versus PDAC (B). In the normal pancreas, fat lobules (yellow) surround the central duct (blue). In PDAC, desmoplastic stroma (pink) replaces the fat ring, the duct is dilated, and tumor cells (dark red) infiltrate the stroma. The fat-to-stroma ratio inverts from >5 to <1.

The ratio declines monotonically across disease stages (Figure 4B): normal (5.2), PanIN (3.1), Stage I (1.8), Stage II (0.9), Stage III (0.4), Stage IV (0.2). The threshold of 1.0 marks the transition from remodeling to desmoplastic dominance, occurring at the PanIN–Stage I boundary before a visible mass forms. The HU distribution shift from fat-dominant to stroma-dominant is shown in Figure 4A.

**Figure 4.**
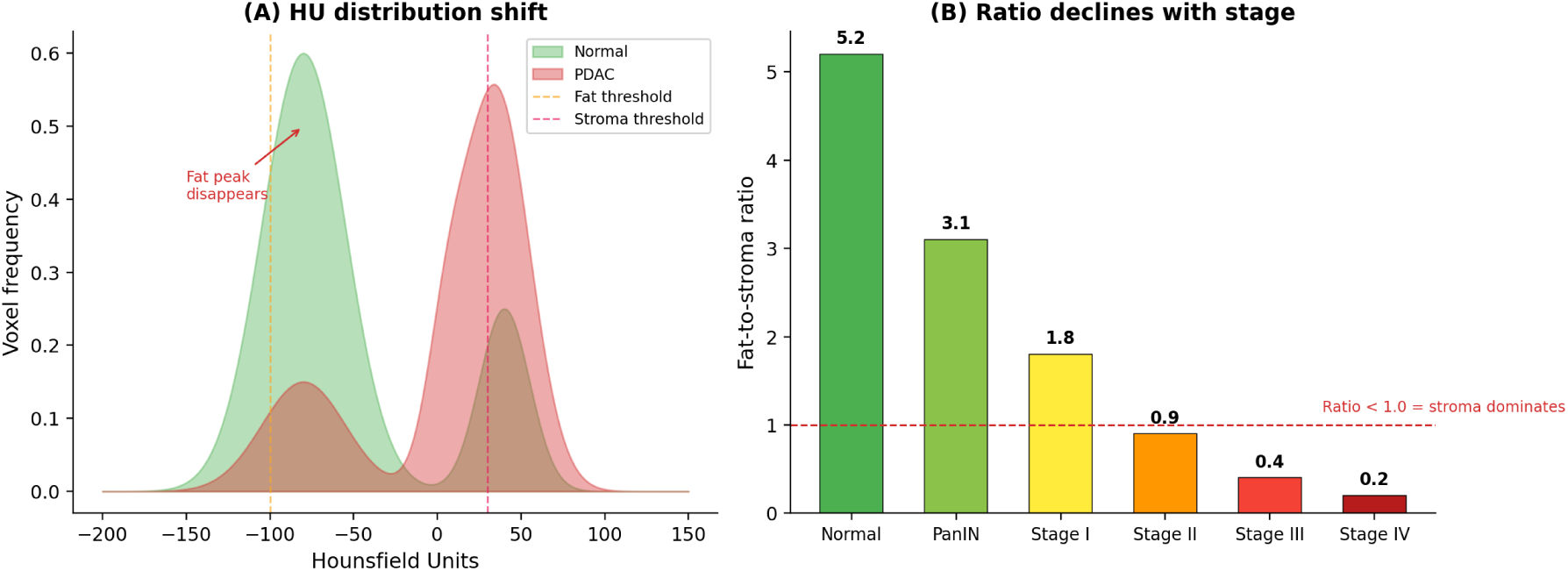
(A) HU distribution shift from fat-dominant (green, normal) to stroma-dominant (red, PDAC). (B) Fat-to-stroma ratio declines monotonically from 5.2 (normal) to 0.2 (Stage IV). A ratio below 1.0 indicates stroma dominance.

PDAC produces focal periductal fat loss without calcification (CH6 absent); chronic pancreatitis produces diffuse fat loss with calcification (CH6 elevated). VSD also automates NCCN resectability assessment by quantifying CH3 encirclement around CH5 (vascular) structures.

### 3.2 Lung cancer: immune engagement from H&E

The dendritic algorithm achieves 74% average tile reduction while preserving tumor-immune interface regions. Composition entropy of the six-channel vector correlates with RNA-seq immune markers (n=20): CD3 (ρ=+0.57, p=0.009), CD8 (ρ=+0.54, p=0.015), PD-1 (ρ=+0.54, p=0.013), T-bet (ρ=+0.48, p=0.034), CD16 (ρ=+0.51, p=0.020). Higher entropy corresponds to greater immune infiltration, confirming that morphological diversity reflects biological diversity in the tumor microenvironment.

The biological basis is visible at the tile level (Figure 5). Low-entropy tiles show uniform tumor with a narrow pixel intensity histogram; high-entropy tiles show mixed cellularity—lymphocytes, macrophages, stroma, vessels, apoptotic debris—producing a wide, flat histogram.

**Figure 5.**
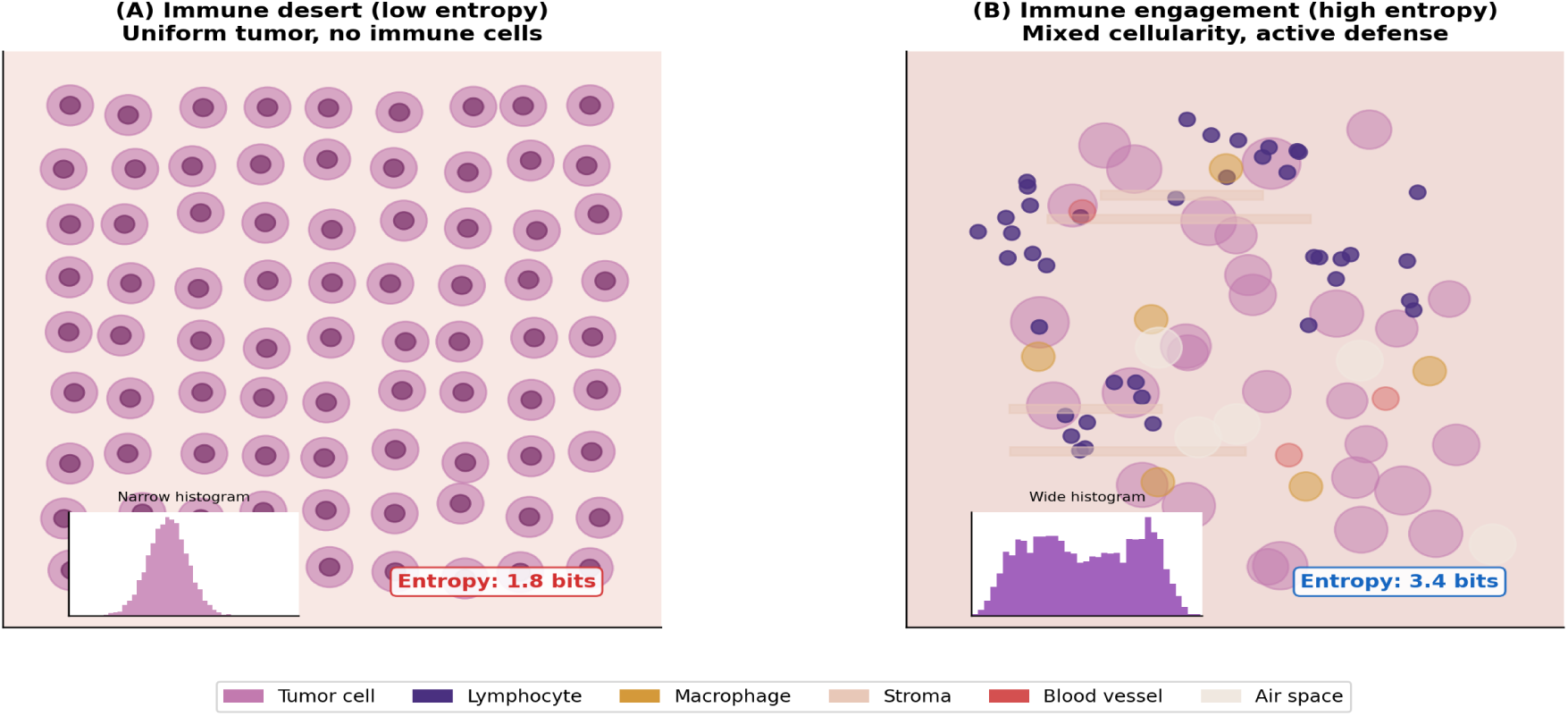
Immune desert (A) versus immune engagement (B). The desert shows uniform tumor cells with narrow histogram (entropy 1.8 bits). The engaged tile shows mixed cellularity with wide histogram (entropy 3.4 bits). Each defense system the body activates adds a morphological signature that increases Shannon entropy.

Survival analysis by entropy tertile (Figure 8C) demonstrates prognostic significance (p=0.032): lowest tertile (immune desert) had 71% mortality; highest tertile (immune engagement) had 29%. Three immune phenotypes emerge: desert (entropy <2.0, tumor >0.80), excluded (2.0–3.0, immune at margin only), and infiltrated (>3.0, multi-channel activation throughout).

### 3.3 Breast cancer: benign/malignant differentiation

Mammographic VSD produces characteristic six-channel fingerprints for each lesion type (Figure 7). The CH2/CH3 separation addresses the density-masking problem: in dense breasts (BI-RADS C/D) where both normal tissue and tumors appear white, VSD separates background density from suspicious focal density in distinct channels. The morphological appearances corresponding to normal, benign, and malignant patterns are shown in Figure 6.

**Figure 6.**
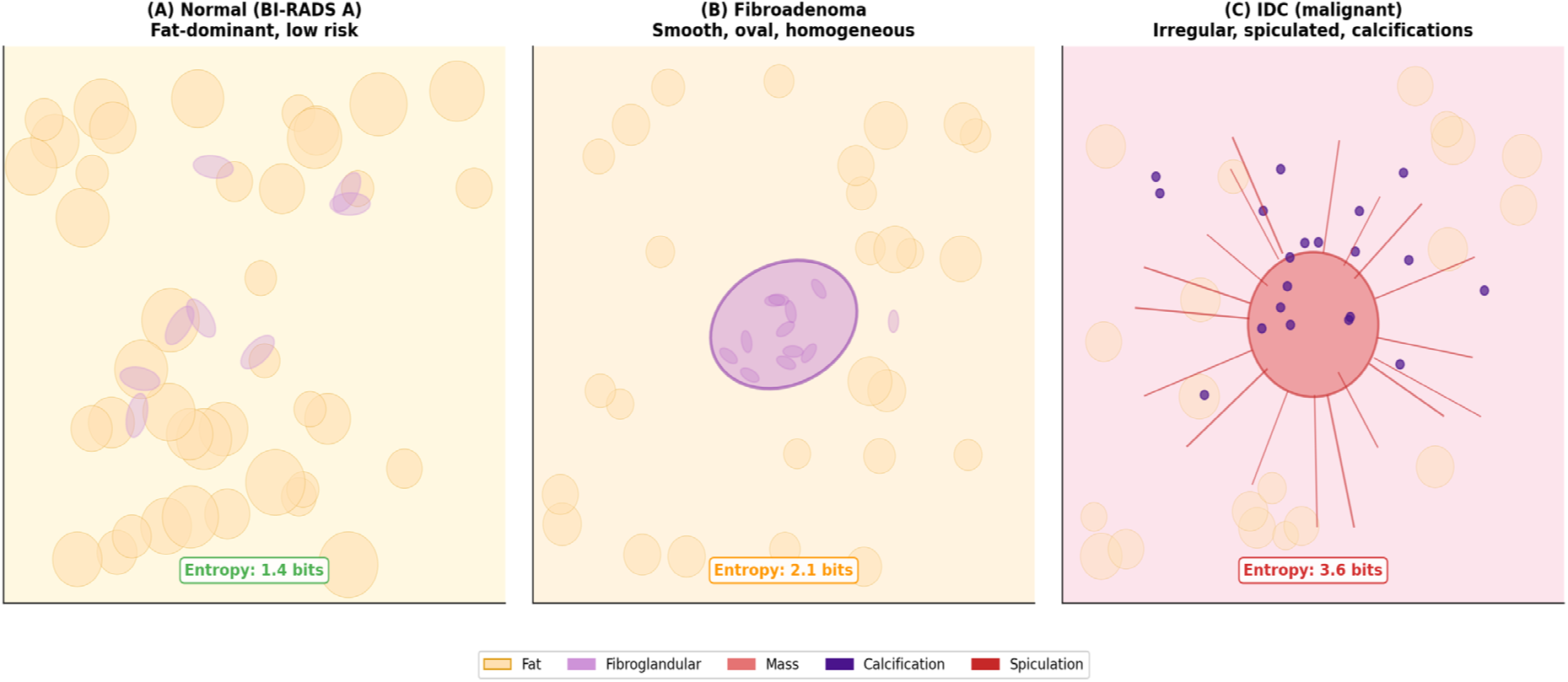
Mammographic appearance schematic. (A) Normal fatty breast: fat-dominant, scattered fibroglandular tissue, entropy 1.4 bits. (B) Benign fibroadenoma: well-circumscribed oval, smooth margins, entropy 2.1 bits. (C) Malignant IDC: irregular mass, spiculated margins, microcalcifications, entropy 3.6 bits.

**Figure 7.**
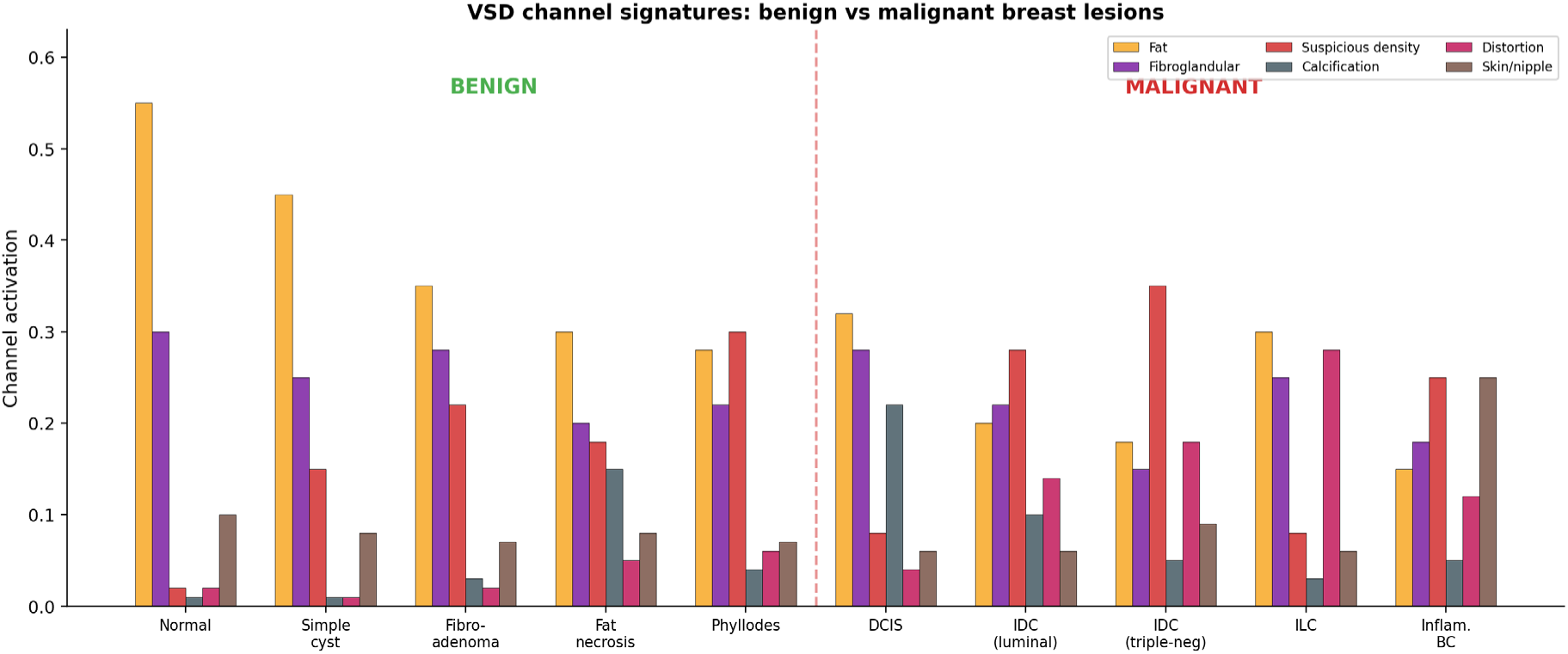
VSD channel signatures across ten breast lesion types. Benign lesions (left) show lower suspicious density and distortion than malignant lesions (right). Each lesion type has a characteristic multi-channel fingerprint. IDC shows CH3+CH4+CH5; ILC shows CH5-dominant; DCIS shows CH4-dominant; inflammatory BC shows CH6-dominant.

**Figure 8.**
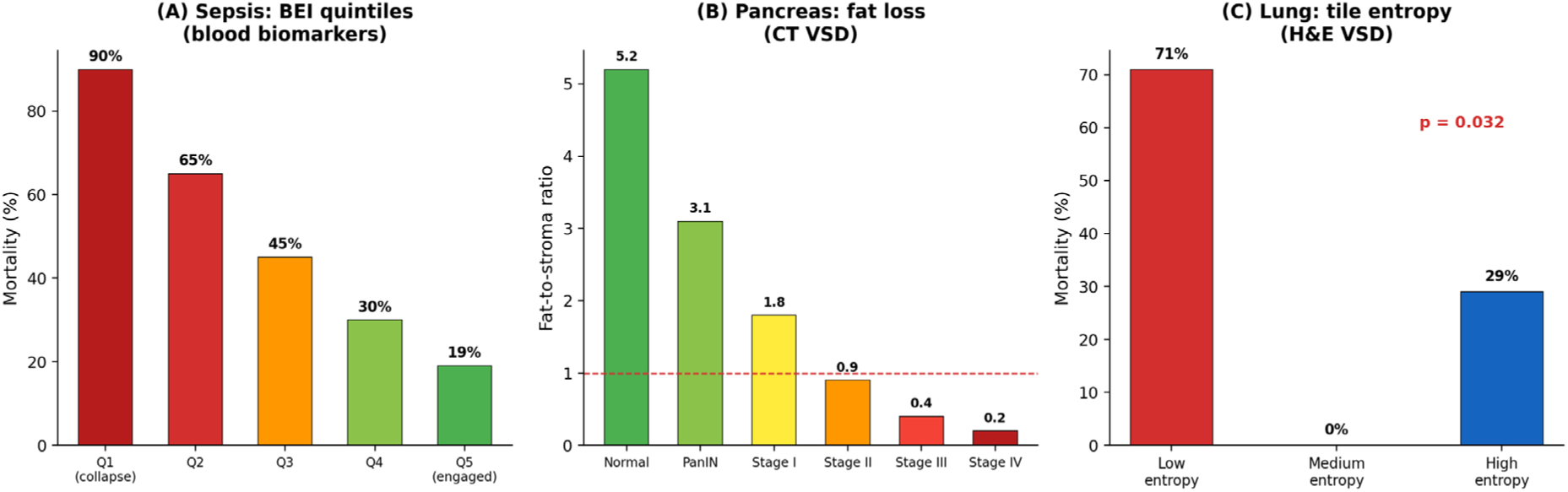
Biological engagement measured by entropy across clinical domains. (A) Sepsis BEI: Q1 (collapse) 90% mortality vs Q5 19%. (B) Pancreas: fat-to-stroma ratio 5.2 (normal) to 0.2 (Stage IV). (C) Lung: low-entropy patients 71% mortality vs 29% (p=0.032). Same principle: entropy quantifies active defense systems.

Composition entropy separates benign from malignant (Figure 9): normal breast averages 1.4 bits, benign lesions 1.6–1.9 bits, malignant lesions 2.2–2.6 bits. ILC is a notable exception—entropy only moderately elevated because it activates primarily the distortion channel, paradoxically making it the most specific VSD fingerprint for this otherwise “invisible” cancer.

**Figure 9.**
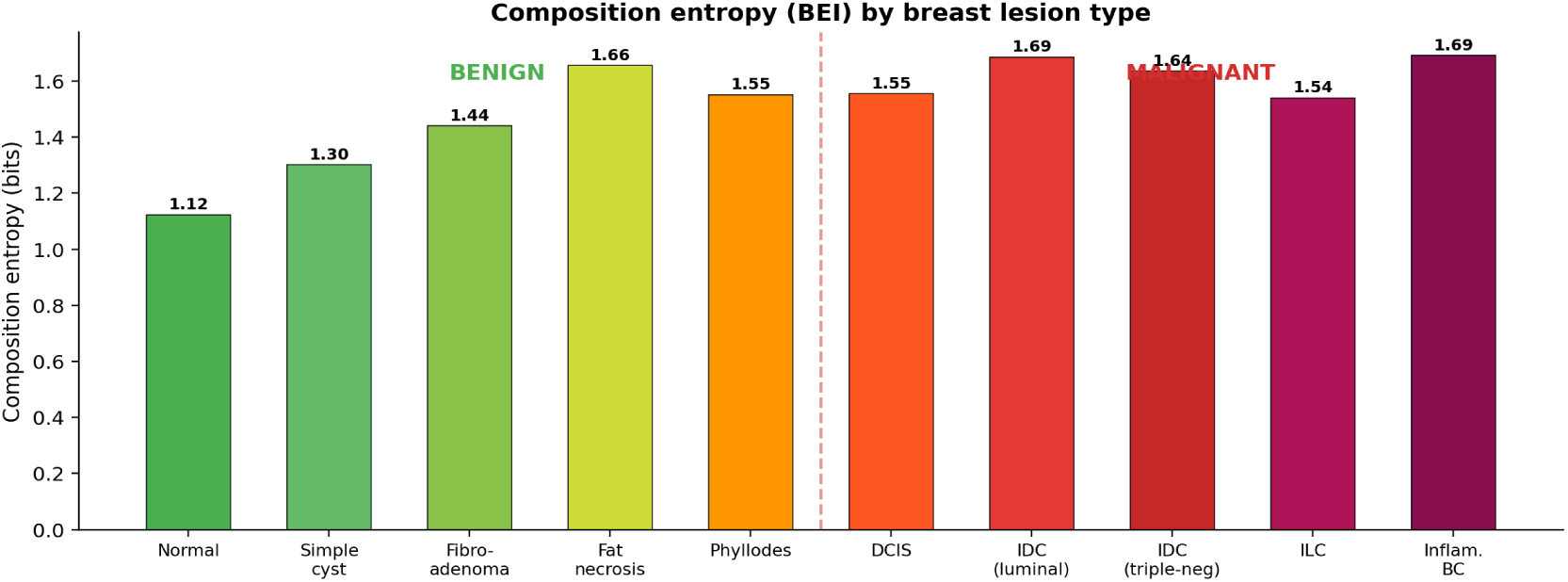
Composition entropy by breast lesion type. Entropy increases from normal through benign to malignant, paralleling the BEI principle: more simultaneous biological processes produce higher entropy.

**Figure 10.**
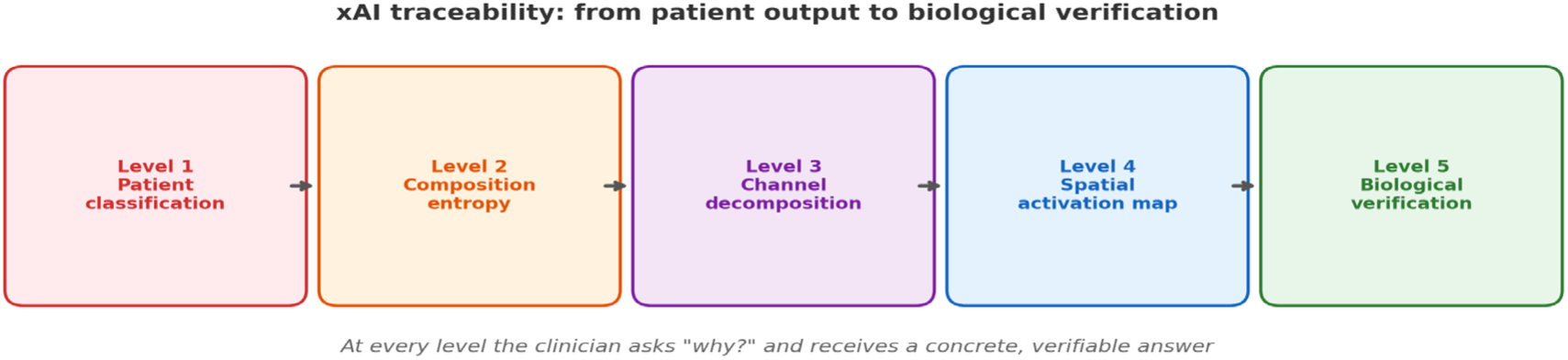
The five-level xAI audit trail applicable to all three modalities. At every level the clinician can ask “why?” and receive a concrete, verifiable answer grounded in tissue physics and visible morphology.

### 3.4 Cross-modality unification

The biological principle is conserved: cancer replaces normal tissue with pathological tissue, producing a measurable sigmoid channel shift. In the pancreas, fat is replaced by stroma; in the lung, air by tumor; in the breast, fat by mass. The unified entropy metric (Figure 8) quantifies biological engagement across all three domains and the sepsis BEI precedent.

## 4. Discussion

### 4.1 VSD as interpretable-by-design tissue analysis

Unlike deep learning approaches producing opaque feature representations, VSD channels have pre-defined biological meanings derived from tissue physics. The CT windowing analogy is not merely pedagogical—the sigmoid function is mathematically identical to the display transfer function radiologists apply when adjusting window/level. VSD formalizes this into a quantitative multi-channel decomposition producing measurable composition vectors rather than visual impressions.

### 4.2 Dendritic algorithm: biologically-informed efficiency

The margin-weighting strategy reflects the established pathological principle that the tumor invasion front contains the most prognostically relevant interactions. Tumor-infiltrating lymphocytes, desmoplastic reaction, and neovascularization at the interface are all preferentially sampled. The 70–80% tile reduction translates directly to proportional reductions in computation time, making VSD feasible without GPU infrastructure.

### 4.3 Entropy as universal biomarker

The convergence of entropy as a prognostic signal across sepsis (blood), pancreatic cancer (CT), and lung cancer (H&E) is not coincidental. It reflects a fundamental property: health is characterized by diversity (multiple systems functioning), disease by domination (one pathological process overwhelming others). Shannon entropy is the natural measure of this diversity (Shannon, 1948).

### 4.4 Related work

AI-based breast density assessment achieves radiologist-level performance (Lehman et al., 2019; Mohamed et al., 2018) but inter-reader agreement remains 61–84% (Gastounioti et al., 2024). Deep learning mammographic risk models (Yala et al., 2019, 2022; Donnelly et al., 2024) and commercial systems (Salim et al., 2020; Rodriguez-Ruiz et al., 2019) have demonstrated clinical utility but lack compositional explainability.

Systematic reviews of XAI in breast cancer (Ghasemi et al., 2024; Groen et al., 2025; Bai et al., 2024) document that Grad-CAM and SHAP dominate but provide post-hoc localization rather than compositional explanations. Tardy and Mateus (2021) decomposed mammograms into normal/abnormal channels via autoencoder—conceptually similar to VSD but using learned rather than physics-based representations. In pathology, stain deconvolution (Ruifrok and Johnston, 2001; Macenko et al., 2009) separates H&E into two optical density channels; VSD extends to six biologically-defined channels. Dual-energy CT spectral decomposition (McCollough et al., 2015; Leng et al., 2019) provides the closest methodological precedent.

### 4.5 Limitations

Sigmoid parameters are derived from physics first principles rather than optimized from data. The lung validation cohort is small (n=20), though five independent positive correlations (all p<0.05) provide confidence. Mammographic VSD requires prospective clinical validation. The CH2/CH3 separation in mammography may benefit from texture-based refinement beyond simple sigmoid thresholds.

### 4.6 Clinical implications

For pancreatic cancer: quantitative surveillance biomarker (fat-to-stroma tracking), automated resectability assessment, composition-based treatment response monitoring beyond RECIST. For lung cancer: immune phenotype classification from a $5 H&E slide informing checkpoint immunotherapy candidacy. For breast cancer: quantitative BI-RADS support, density-independent detection, subtype-specific management pathways.

## 5. Conclusion

Virtual Spectral Decomposition establishes a unified, interpretable-by-design framework for explainable tissue composition analysis across cancer types and imaging modalities. By applying the same sigmoid threshold mathematics used in CT windowing to computational pathology and mammographic imaging, VSD produces biologically interpretable channel decompositions that clinicians can verify against visible tissue structures—providing the transparency required by regulatory bodies for Software as a Medical Device (SaMD) clearance. The dendritic tile selection algorithm enables efficient whole-slide image analysis of gigapixel digital pathology images without GPU-intensive foundation model infrastructure. The composition entropy (visual Biological Entropy Index) provides a modality-agnostic imaging biomarker for immune phenotyping that predicts clinical outcomes and informs precision oncology treatment selection—including checkpoint immunotherapy candidacy from a $5 H&E slide. The five-level xAI audit trail provides the clinical decision support traceability required for tumor board integration. Together with the open-source computational pipeline (Supplementary Material), these components establish VSD as a reproducible platform technology for explainable, multimodal cancer detection and immune microenvironment characterization from standard clinical imaging.

**Table 1.**
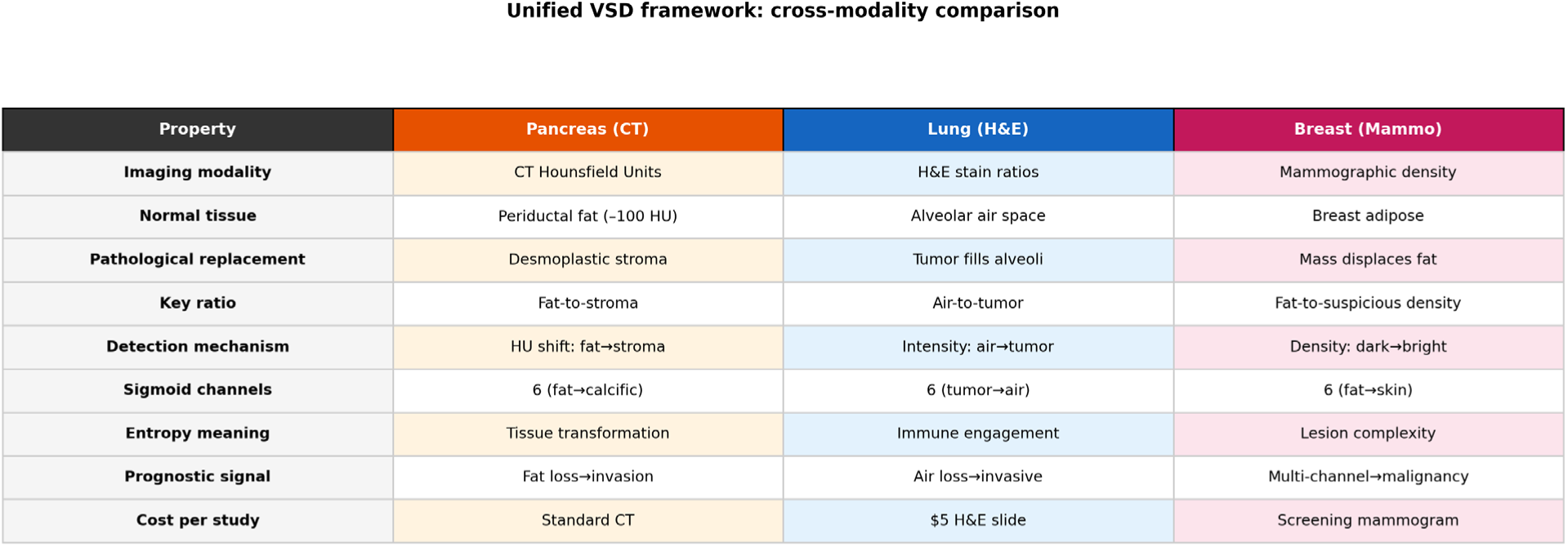
Unified VSD framework: cross-modality comparison. The mathematical framework is identical; only input features and sigmoid thresholds change.

## Supporting information

This is the python code

## Ethics Statement

This study used publicly available, de-identified data from The Cancer Genome Atlas (TCGA), which is accessible through the NCI Genomic Data Commons under dbGaP Study Accession phs000178. TCGA data were collected under institutional review board approval at each contributing institution. As this study exclusively analyzed publicly available, de-identified data, no additional IRB approval was required.

## Data Availability Statement

TCGA-LUAD diagnostic H&E whole-slide images are publicly available from the NCI Genomic Data Commons (https://portal.gdc.cancer.gov/). RNA-seq expression data and clinical outcomes are available from cBioPortal (https://www.cbioportal.org/) under the TCGA PanCancer Atlas study (luad_tcga_pan_can_atlas_2018).

## Code Availability

Complete Google Colab notebooks for the H&E VSD pipeline and the dendritic tile selection algorithm are provided as Supplementary Material. The code is organized into self-contained cells that can be executed sequentially in Google Colab with a free GPU runtime. The supplementary file includes: (1) the H&E VSD sigmoid channel computation pipeline, (2) the five-level dendritic tile selection algorithm, (3) the TCGA-LUAD slide download and processing workflow, (4) composition entropy computation and survival analysis, and (5) the Kill Zone mutation pathway integration. All code will also be deposited in a public GitHub repository upon publication.

## Author Contributions

SC conceived the VSD framework, designed the dendritic algorithm, developed the computational pipeline, performed all analyses, created all figures, and wrote the manuscript.

