## Supplementary material for "Virtual Spectral Decomposition with Dendritic Tile Selection: An Explainable AI Framework for Multimodal Tissue Composition Analysis and Immune Phenotyping Across Pancreatic, Lung, and Breast Cancer": This is the python code: Supplementary_Code_VSD_Pipeline.py

```
# ══════════════════════════════════════════════════════════════════════════════
# SUPPLEMENTARY CODE — medRxiv Preprint
#
# Virtual Spectral Decomposition with Dendritic Tile Selection:
# A Unified, Explainable Framework for Tissue Composition Analysis
# Across Pancreatic, Lung, and Breast Cancer
#
# Shubham Chandra, Chandra Associates, Dallas, Texas
# April 2026
#
# USAGE: Copy each cell (separated by # ═══ CELL N ═══) into a Google Colab
# notebook and execute sequentially. Requires GPU runtime for feature
# extraction (Runtime > Change runtime type > GPU).
#
# OUTPUTS: All results saved to Google Drive at:
#   /content/drive/MyDrive/VSD_Output/
# ══════════════════════════════════════════════════════════════════════════════


# ══════════════════════════════════════════════════════════════════════════════
# CELL 1: Environment Setup
# ══════════════════════════════════════════════════════════════════════════════

# --- Run these lines in a Colab code cell ---
# !pip install openslide-python lifelines tqdm
# !apt-get install -y openslide-tools

import os, json, time, warnings
import numpy as np
import pandas as pd
import matplotlib.pyplot as plt
from PIL import Image
from scipy import ndimage
from scipy.stats import entropy, spearmanr, mannwhitneyu
from sklearn.preprocessing import StandardScaler
from sklearn.ensemble import RandomForestClassifier
from sklearn.metrics import roc_auc_score
from sklearn.model_selection import StratifiedKFold
from lifelines import CoxPHFitter, KaplanMeierFitter
from lifelines.statistics import logrank_test
from tqdm import tqdm
import requests
import openslide

warnings.filterwarnings('ignore')

# --- Mount Google Drive ---
# from google.colab import drive
# drive.mount('/content/drive')

# --- Output directories ---
OUTPUT_DIR = '/content/drive/MyDrive/VSD_Output'
SLIDE_DIR  = '/content/slides_local'   # Local disk for slides (no Drive quota issues)
FIG_DIR    = os.path.join(OUTPUT_DIR, 'figures')
DATA_DIR   = os.path.join(OUTPUT_DIR, 'data')
for d in [OUTPUT_DIR, SLIDE_DIR, FIG_DIR, DATA_DIR]:
    os.makedirs(d, exist_ok=True)

print(f'Output directory: {OUTPUT_DIR}')
print(f'Slide directory:  {SLIDE_DIR} (local disk)')
print('Environment ready.')


# ══════════════════════════════════════════════════════════════════════════════
# CELL 2: H&E VSD — Sigmoid Channel Definitions
# ══════════════════════════════════════════════════════════════════════════════
#
# Implements the six-channel sigmoid decomposition described in Section 2.3.2
# of the manuscript. Each channel is a sigmoid function applied to a derived
# staining feature, analogous to CT HU thresholds in pancreatic VSD.
#
# Channel mapping:
#   CH1: Tumor cellularity    — high hematoxylin + high tissue intensity
#   CH2: Stromal desmoplasia  — high eosin, low nuclear density
#   CH3: Immune infiltrate    — nuclear size heterogeneity (variance)
#   CH4: Necrosis / apoptosis — bimodal intensity distribution
#   CH5: Vascular             — red channel excess
#   CH6: Air space            — very bright pixels (alveolar air)
# ══════════════════════════════════════════════════════════════════════════════

def sigmoid(x, center, width):
    """Sigmoid activation — identical to CT windowing transfer function.
    σ(x; c, w) = 1 / (1 + exp(-(x - c) / w))
    """
    z = -(x - center) / (width + 1e-8)
    return 1.0 / (1.0 + np.exp(np.clip(z, -30, 30)))


def rgb_to_stain_features(tile_rgb):
    """Convert RGB tile to stain-space features derived from H&E physics.
    
    Hematoxylin (blue/purple) binds nucleic acids → marks nuclei.
    Eosin (pink) binds proteins → marks cytoplasm, stroma, collagen.
    """
    arr = tile_rgb.astype(np.float32) / 255.0
    r, g, b = arr[:,:,0], arr[:,:,1], arr[:,:,2]
    
    nuclear = b / (r + 1e-4)                    # Hematoxylin/eosin ratio
    stromal = (r + g) / (2.0 * b + 1e-4)        # Eosin dominance
    intensity = 1.0 - (r + g + b) / 3.0         # Inverted brightness
    red_excess = r / (g + b + 1e-4)              # Blood vessel indicator
    
    from scipy.ndimage import uniform_filter
    nuc_mean = uniform_filter(nuclear, size=16)
    nuc_sq_mean = uniform_filter(nuclear**2, size=16)
    nuc_var = np.clip(nuc_sq_mean - nuc_mean**2, 0, None)
    
    return {'nuclear': nuclear, 'stromal': stromal, 'intensity': intensity,
            'red_excess': red_excess, 'nuc_var': nuc_var}


def compute_vsd_channels(tile_rgb):
    """Compute 6 sigmoid VSD channels for one H&E tile (256x256).
    Returns dict of channel name → mean activation (0-1).
    """
    feats = rgb_to_stain_features(tile_rgb)
    channels = {}
    
    # CH1: Tumor cellularity
    ch_nuc = sigmoid(feats['nuclear'], center=0.95, width=0.08)
    ch_int = sigmoid(feats['intensity'], center=0.35, width=0.10)
    channels['tumor_cellularity'] = float(np.mean(ch_nuc * ch_int))
    
    # CH2: Stromal desmoplasia (lung analog of pancreatic fat loss)
    ch_stroma = sigmoid(feats['stromal'], center=1.3, width=0.15)
    ch_low_nuc = 1.0 - sigmoid(feats['nuclear'], center=0.85, width=0.10)
    channels['stromal_desmoplasia'] = float(np.mean(ch_stroma * ch_low_nuc))
    
    # CH3: Immune infiltrate (nuclear size heterogeneity)
    ch_hetero = sigmoid(feats['nuc_var'], center=0.02, width=0.008)
    ch_tissue = sigmoid(feats['intensity'], center=0.20, width=0.10)
    channels['immune_infiltrate'] = float(np.mean(ch_hetero * ch_tissue))
    
    # CH4: Necrosis / apoptosis (bimodal intensity)
    dark = sigmoid(feats['intensity'], center=0.60, width=0.08)
    pale = sigmoid(feats['intensity'], center=0.12, width=0.05)
    channels['necrosis_apoptosis'] = float(min(np.mean(dark) * np.mean(pale) * 4, 1.0))
    
    # CH5: Vascular (red channel excess)
    ch_red = sigmoid(feats['red_excess'], center=0.85, width=0.10)
    channels['vascular'] = float(np.mean(ch_red * ch_tissue))
    
    # CH6: Air space (bright pixels = alveolar air)
    bright = 1.0 - feats['intensity']
    ch_air = sigmoid(bright, center=0.75, width=0.08)
    channels['air_space'] = float(np.mean(ch_air))
    
    # Composition entropy (visual Biological Entropy Index)
    vec = np.array(list(channels.values()))
    vec_norm = vec / (vec.sum() + 1e-8)
    channels['composition_entropy'] = float(entropy(vec_norm + 1e-10))
    
    return channels

VSD_CHANNELS = ['tumor_cellularity', 'stromal_desmoplasia', 'immune_infiltrate',
                'necrosis_apoptosis', 'vascular', 'air_space']

print('H&E VSD channels defined (6 sigmoid channels + composition entropy)')


# ══════════════════════════════════════════════════════════════════════════════
# CELL 3: Dendritic Tile Selection Algorithm (Section 2.2)
# ══════════════════════════════════════════════════════════════════════════════
#
# Five-level hierarchical processing:
#   Level 0 (Soma):      Tissue mask via saturation thresholding
#   Level 1 (Primary):   Coarse patch screening at 8x downsample
#   Level 2 (Secondary): Entropy-based subpatch selection + margin detection
#   Level 3 (Terminal):  Full-resolution tile reading + VSD computation
#   Level 4 (Integration): Spatially-weighted aggregation
# ══════════════════════════════════════════════════════════════════════════════

CONFIG = {
    'thumbnail_max': 1024,
    'tissue_sat_thresh': 0.15,
    'tile_size': 256,
    'downsample_screen': 4,
    'entropy_percentile': 70,
    'max_tiles': 500,
    'min_tissue_frac': 0.40,
    'blank_thresh': 240,
    'std_thresh': 10,
}


def get_tissue_mask(slide):
    """Level 0: Soma — identify tissue vs background."""
    dims = slide.level_dimensions[0]
    scale = min(CONFIG['thumbnail_max'] / dims[0], CONFIG['thumbnail_max'] / dims[1])
    w, h = int(dims[0] * scale), int(dims[1] * scale)
    thumb = np.array(slide.get_thumbnail((w, h)).convert('RGB')).astype(float) / 255.0
    r, g, b = thumb[:,:,0], thumb[:,:,1], thumb[:,:,2]
    cmax = np.maximum(np.maximum(r, g), b)
    cmin = np.minimum(np.minimum(r, g), b)
    sat = np.where(cmax > 0, (cmax - cmin) / (cmax + 1e-8), 0)
    mask = sat > CONFIG['tissue_sat_thresh']
    mask = ndimage.binary_fill_holes(mask)
    mask = ndimage.binary_opening(mask, iterations=2)
    mask = ndimage.binary_closing(mask, iterations=2)
    return mask, scale, dims


def select_tiles_dendritic(slide, mask, scale, dims):
    """Levels 1-2: Primary + Secondary dendrites.
    Returns list of (x, y) coordinates for full-resolution VSD."""
    tile_sz = CONFIG['tile_size']
    ds = CONFIG['downsample_screen']
    step = tile_sz * ds
    
    screen_level = 0
    for lvl in range(len(slide.level_dimensions)):
        if slide.level_downsamples[lvl] <= ds:
            screen_level = lvl
    actual_ds = slide.level_downsamples[screen_level]
    read_sz = int(tile_sz * ds / actual_ds)
    
    candidates = []
    for y in range(0, dims[1] - step, step):
        for x in range(0, dims[0] - step, step):
            tx, ty = int(x * scale), int(y * scale)
            tw = max(1, int(step * scale))
            th = max(1, int(step * scale))
            if ty >= mask.shape[0] or tx >= mask.shape[1]:
                continue
            region = mask[ty:min(ty+th, mask.shape[0]), tx:min(tx+tw, mask.shape[1])]
            if region.size == 0 or region.mean() < CONFIG['min_tissue_frac']:
                continue
            try:
                img = slide.read_region((x, y), screen_level, (read_sz, read_sz)).convert('RGB')
                arr = np.array(img)
                if arr.mean() > CONFIG['blank_thresh'] or arr.std() < CONFIG['std_thresh']:
                    continue
                gray = arr.mean(axis=2).astype(np.uint8)
                hist, _ = np.histogram(gray, bins=64, range=(0, 256))
                hist = hist / (hist.sum() + 1e-8)
                ent = entropy(hist + 1e-10)
                nuc = arr[:,:,2].mean() / (arr[:,:,0].mean() + 1e-8)
                is_margin = 0.3 < nuc < 0.9
                candidates.append({'x': x, 'y': y, 'entropy': ent,
                                   'is_margin': is_margin, 'nuclear': nuc})
            except:
                continue
    
    if not candidates:
        return []
    
    scores = [c['entropy'] for c in candidates]
    thresh = np.percentile(scores, CONFIG['entropy_percentile'])
    selected = [c for c in candidates if c['entropy'] >= thresh]
    margins = [c for c in candidates if c['is_margin'] and c not in selected]
    selected.extend(margins[:max(50, len(selected) // 4)])
    selected.sort(key=lambda c: c['entropy'], reverse=True)
    return selected[:CONFIG['max_tiles']]


def process_slide_vsd(wsi_path):
    """Full VSD pipeline: Levels 0-4 for one slide.
    Returns per-slide VSD composition vector + metadata."""
    slide = openslide.OpenSlide(wsi_path)
    name = os.path.basename(wsi_path)
    dims = slide.level_dimensions[0]
    print(f'\n  [{name}] {dims[0]}\u00d7{dims[1]}')
    
    t0 = time.time()
    mask, scale, dims = get_tissue_mask(slide)
    tissue_frac = mask.mean()
    print(f'    Tissue: {tissue_frac*100:.1f}%')
    if tissue_frac < 0.05:
        slide.close(); return None
    
    tiles = select_tiles_dendritic(slide, mask, scale, dims)
    print(f'    Selected: {len(tiles)} tiles')
    if len(tiles) < 5:
        slide.close(); return None
    
    # Level 3: Full-resolution VSD
    tile_sz = CONFIG['tile_size']
    tile_vsd, tile_meta = [], []
    for t in tiles:
        try:
            img = slide.read_region((t['x'], t['y']), 0, (tile_sz, tile_sz)).convert('RGB')
            arr = np.array(img)
            if arr.mean() > CONFIG['blank_thresh'] or arr.std() < CONFIG['std_thresh']:
                continue
            vsd = compute_vsd_channels(arr)
            tile_vsd.append(vsd)
            tile_meta.append(t)
        except:
            continue
    
    slide.close()
    if len(tile_vsd) < 5:
        return None
    
    elapsed = time.time() - t0
    print(f'    VSD: {len(tile_vsd)} tiles in {elapsed:.1f}s')
    
    # Level 4: Dendritic integration (spatially-weighted)
    weights = np.ones(len(tile_meta))
    median_ent = np.median([m['entropy'] for m in tile_meta])
    for i, m in enumerate(tile_meta):
        if m['is_margin']:    weights[i] *= 2.0
        if m['entropy'] > median_ent: weights[i] *= 1.5
    weights /= weights.sum()
    
    all_channels = VSD_CHANNELS + ['composition_entropy']
    slide_vsd = {}
    for ch in all_channels:
        vals = np.array([tv[ch] for tv in tile_vsd])
        slide_vsd[f'vsd_{ch}_mean'] = float(np.average(vals, weights=weights))
        slide_vsd[f'vsd_{ch}_std'] = float(np.sqrt(
            np.average((vals - np.average(vals, weights=weights))**2, weights=weights)))
    
    air_vals = np.array([tv['air_space'] for tv in tile_vsd])
    tumor_vals = np.array([tv['tumor_cellularity'] for tv in tile_vsd])
    
    return {
        **slide_vsd,
        'margin_frac': float(np.mean([m['is_margin'] for m in tile_meta])),
        'mean_tile_entropy': float(np.mean([m['entropy'] for m in tile_meta])),
        'air_tumor_ratio': float(np.mean(air_vals) / (np.mean(tumor_vals) + 1e-8)),
        'stroma_invasion_score': float(np.mean([tv['stromal_desmoplasia'] for tv in tile_vsd])
                                       / (np.mean(air_vals) + 1e-8)),
        'n_tiles': len(tile_vsd),
        'n_candidates': len(tiles),
        'reduction_pct': 1.0 - len(tile_vsd) / max(len(tiles), 1),
        'time_s': elapsed,
    }

print('Dendritic VSD pipeline defined.')


# ══════════════════════════════════════════════════════════════════════════════
# CELL 4: Download TCGA-LUAD Slides from GDC (to local disk)
# ══════════════════════════════════════════════════════════════════════════════
#
# Downloads diagnostic H&E slides directly to Colab local disk (/content/).
# This avoids Google Drive quota issues that cause I/O errors.
# Adjust MAX_SLIDES to control how many slides to process.
# ══════════════════════════════════════════════════════════════════════════════

MAX_SLIDES = 10  # Start small; increase to 500 once pipeline is verified

print(f'Querying GDC for TCGA-LUAD diagnostic slides (max {MAX_SLIDES})...')

filters = {
    "op": "and",
    "content": [
        {"op": "in", "content": {"field": "cases.project.project_id", "value": ["TCGA-LUAD"]}},
        {"op": "in", "content": {"field": "data_type", "value": ["Slide Image"]}},
        {"op": "in", "content": {"field": "experimental_strategy", "value": ["Diagnostic Slide"]}},
    ]
}
params = {
    "filters": json.dumps(filters),
    "fields": "file_id,file_name,file_size,cases.submitter_id",
    "format": "JSON",
    "size": MAX_SLIDES,
}

r = requests.get('https://api.gdc.cancer.gov/files', params=params)
hits = r.json()['data']['hits']
print(f'Found {len(hits)} slides')

downloaded = []
for hit in tqdm(hits, desc='Downloading'):
    fid = hit['file_id']
    fname = hit['file_name']
    dst = os.path.join(SLIDE_DIR, fname)
    patient = hit.get('cases', [{}])[0].get('submitter_id', fname[:12])
    
    if os.path.exists(dst) and os.path.getsize(dst) > 1e6:
        print(f'  Already have: {fname}')
        downloaded.append({'file_path': dst, 'file_name': fname,
                           'patient_id': patient, 'file_id': fid})
        continue
    
    try:
        print(f'  Downloading {fname}...', end=' ', flush=True)
        resp = requests.get(f'https://api.gdc.cancer.gov/data/{fid}', stream=True, timeout=600)
        with open(dst, 'wb') as f:
            for chunk in resp.iter_content(65536):
                f.write(chunk)
        sz = os.path.getsize(dst)
        print(f'{sz/1e6:.0f} MB')
        # Verify OpenSlide can open it
        slide = openslide.OpenSlide(dst)
        slide.close()
        downloaded.append({'file_path': dst, 'file_name': fname,
                           'patient_id': patient, 'file_id': fid})
    except Exception as e:
        print(f'FAILED: {e}')

print(f'\nDownloaded {len(downloaded)} slides to {SLIDE_DIR}')


# ══════════════════════════════════════════════════════════════════════════════
# CELL 5: Run VSD Pipeline on All Slides
# ══════════════════════════════════════════════════════════════════════════════

print('=' * 70)
print('RUNNING H&E VSD PIPELINE')
print('=' * 70)

vsd_results = []
for slide_info in tqdm(downloaded, desc='VSD'):
    try:
        result = process_slide_vsd(slide_info['file_path'])
        if result is None:
            continue
        result['file_name'] = slide_info['file_name']
        result['patient_id'] = slide_info['patient_id']
        result['sampleId'] = slide_info['file_name'].split('.')[0][:15]
        vsd_results.append(result)
    except Exception as e:
        print(f'  ERROR: {slide_info["file_name"]}: {e}')

print(f'\n{"="*70}')
print(f'VSD COMPLETE: {len(vsd_results)}/{len(downloaded)} slides')
print(f'{"="*70}')

vsd_df = pd.DataFrame(vsd_results)
vsd_df.to_csv(os.path.join(DATA_DIR, 'hne_vsd_results.csv'), index=False)
print(f'Saved: {DATA_DIR}/hne_vsd_results.csv')


# ══════════════════════════════════════════════════════════════════════════════
# CELL 6: VSD Channel Summary and Visualization
# ══════════════════════════════════════════════════════════════════════════════

if len(vsd_df) > 0:
    print('\n── H&E VSD Channel Summary ──')
    for ch in VSD_CHANNELS:
        col = f'vsd_{ch}_mean'
        if col in vsd_df.columns:
            print(f'  {ch:<25}: {vsd_df[col].mean():.4f} \u00b1 {vsd_df[col].std():.4f}')
    
    print(f'  composition_entropy      : {vsd_df["vsd_composition_entropy_mean"].mean():.4f}')
    print(f'  air_tumor_ratio          : {vsd_df["air_tumor_ratio"].mean():.4f}')
    print(f'  stroma_invasion_score    : {vsd_df["stroma_invasion_score"].mean():.4f}')
    
    # Channel distributions
    fig, axes = plt.subplots(2, 3, figsize=(15, 9))
    colors = ['#D32F2F', '#E91E63', '#7B1FA2', '#455A64', '#C62828', '#1565C0']
    for i, ch in enumerate(VSD_CHANNELS):
        ax = axes[i//3, i%3]
        col = f'vsd_{ch}_mean'
        vals = vsd_df[col].dropna()
        ax.hist(vals, bins=25, color=colors[i], edgecolor='black', alpha=0.8)
        ax.set_title(f'CH{i+1}: {ch}', fontweight='bold')
        ax.set_xlabel('Mean activation'); ax.set_ylabel('Count')
        ax.axvline(vals.mean(), color='black', linestyle='--', linewidth=1.5)
        ax.spines['top'].set_visible(False); ax.spines['right'].set_visible(False)
    plt.suptitle('H&E VSD Channel Distributions', fontsize=14, fontweight='bold', y=1.02)
    plt.tight_layout()
    plt.savefig(os.path.join(FIG_DIR, 'vsd_channel_distributions.png'), dpi=150, bbox_inches='tight')
    plt.show()
    
    # Air-tumor ratio vs stroma invasion
    fig, ax = plt.subplots(figsize=(8, 6))
    sc = ax.scatter(vsd_df['air_tumor_ratio'], vsd_df['stroma_invasion_score'],
                    c=vsd_df['vsd_composition_entropy_mean'], cmap='RdYlBu_r',
                    s=60, edgecolor='black', linewidth=0.5, alpha=0.8)
    ax.set_xlabel('Air-to-tumor ratio (high = preserved alveoli)')
    ax.set_ylabel('Stroma invasion score (high = desmoplasia replacing air)')
    ax.set_title('H&E VSD: Air Loss vs Stroma Replacement\n(Color = Composition Entropy)')
    plt.colorbar(sc, label='Composition Entropy')
    ax.spines['top'].set_visible(False); ax.spines['right'].set_visible(False)
    plt.tight_layout()
    plt.savefig(os.path.join(FIG_DIR, 'vsd_air_vs_stroma.png'), dpi=150, bbox_inches='tight')
    plt.show()

print('Visualization complete.')


# ══════════════════════════════════════════════════════════════════════════════
# CELL 7: Merge with Clinical/Survival Data and Survival Analysis
# ══════════════════════════════════════════════════════════════════════════════
#
# Requires: tcga_luad_survival.csv from cBioPortal (TCGA PanCancer Atlas)
# Download from: https://www.cbioportal.org/study/summary?id=luad_tcga_pan_can_atlas_2018
# Place in: /content/drive/MyDrive/VSD_Output/data/
# ══════════════════════════════════════════════════════════════════════════════

SURV_PATH = os.path.join(DATA_DIR, 'tcga_luad_survival.csv')

if os.path.exists(SURV_PATH) and len(vsd_df) > 0:
    surv_df = pd.read_csv(SURV_PATH)
    
    vsd_df['sampleId_match'] = vsd_df['sampleId'].str[:len(surv_df['sampleId'].iloc[0])]
    merged = surv_df.merge(vsd_df, left_on='sampleId', right_on='sampleId_match', how='inner')
    print(f'Merged: {len(merged)} patients with VSD + survival data')
    
    if len(merged) >= 15:
        # Entropy tertile survival analysis
        ent_col = 'vsd_composition_entropy_mean'
        merged['entropy_group'] = pd.qcut(merged[ent_col], q=3,
                                           labels=['Low', 'Medium', 'High'], duplicates='drop')
        
        fig, ax = plt.subplots(figsize=(8, 6))
        kmf = KaplanMeierFitter()
        colors = {'Low': '#D32F2F', 'Medium': '#F9A825', 'High': '#1565C0'}
        
        for grp in ['Low', 'Medium', 'High']:
            g = merged[merged['entropy_group'] == grp]
            if len(g) > 0:
                kmf.fit(g['time'], g['event'], label=f'{grp} entropy (n={len(g)})')
                kmf.plot_survival_function(ax=ax, color=colors[grp], linewidth=2.5, ci_show=True)
        
        low = merged[merged['entropy_group'] == 'Low']
        high = merged[merged['entropy_group'] == 'High']
        if len(low) > 5 and len(high) > 5:
            lr = logrank_test(low['time'], high['time'], low['event'], high['event'])
            ax.set_title(f'H&E VSD Composition Entropy \u2014 Survival\n(p={lr.p_value:.4f})', fontweight='bold')
        
        ax.set_xlabel('Overall Survival (Months)')
        ax.set_ylabel('Survival Probability')
        ax.legend(loc='lower left')
        ax.spines['top'].set_visible(False); ax.spines['right'].set_visible(False)
        plt.tight_layout()
        plt.savefig(os.path.join(FIG_DIR, 'vsd_entropy_survival.png'), dpi=150, bbox_inches='tight')
        plt.show()
        print('Survival analysis complete.')
    else:
        print(f'Only {len(merged)} matched patients. Increase MAX_SLIDES.')
else:
    print(f'Survival file not found at {SURV_PATH}.')
    print('Download from cBioPortal: luad_tcga_pan_can_atlas_2018')


# ══════════════════════════════════════════════════════════════════════════════
# CELL 8: Immune Marker Correlation (RNA-seq validation)
# ══════════════════════════════════════════════════════════════════════════════
#
# Tests correlation between VSD composition entropy and RNA-seq immune markers.
# Requires expression data from cBioPortal.
# ══════════════════════════════════════════════════════════════════════════════

IMMUNE_GENES = {
    'CD3':  ['CD3D', 'CD3E', 'CD3G'],
    'CD8':  ['CD8A', 'CD8B'],
    'PD1':  ['PDCD1'],
    'Tbet': ['TBX21'],
    'CD16': ['FCGR3A'],
    'CD68': ['CD68'],
    'PDL1': ['CD274'],
    'FOXP3':['FOXP3'],
    'Ki67': ['MKI67'],
}

EXPR_PATH = os.path.join(DATA_DIR, 'tcga_luad_expression.csv')

if os.path.exists(EXPR_PATH) and 'merged' in dir() and len(merged) > 0:
    expr_df = pd.read_csv(EXPR_PATH)
    
    # Compute composite immune scores
    for score_name, genes in IMMUNE_GENES.items():
        available = [g for g in genes if g in expr_df.columns]
        if available:
            expr_df[f'{score_name}_score'] = expr_df[available].mean(axis=1)
    
    # Merge with VSD
    full = merged.merge(expr_df, on='sampleId', how='inner')
    
    if len(full) >= 10:
        print(f'\n── Immune Marker Correlations (n={len(full)}) ──')
        ent_col = 'vsd_composition_entropy_mean'
        print(f'{"Marker":<15} {"rho":>8} {"p-value":>10} {"Direction":>10}')
        print('-' * 45)
        
        for score_name in IMMUNE_GENES:
            col = f'{score_name}_score'
            if col in full.columns:
                rho, pval = spearmanr(full[ent_col], full[col])
                direction = '\u2191' if rho > 0 else '\u2193'
                sig = '*' if pval < 0.05 else ''
                print(f'  {score_name:<13} {rho:>8.3f} {pval:>10.4f} {direction:>5} {sig}')
else:
    print('Expression data not found. Skipping immune correlation.')
    print(f'Expected at: {EXPR_PATH}')


# ══════════════════════════════════════════════════════════════════════════════
# CELL 9: Summary Report
# ══════════════════════════════════════════════════════════════════════════════

from datetime import datetime

report = f"""
================================================================================
H&E VIRTUAL SPECTRAL DECOMPOSITION — RESULTS SUMMARY
Generated: {datetime.now().strftime('%Y-%m-%d %H:%M')}
================================================================================

CONCEPT:
  Sigmoid channel decomposition of H&E staining into 6 biological channels.
  Same math as CT windowing. Composition entropy = visual BEI.
  
  Pancreas analog: fat loss around duct -> desmoplasia -> cancer
  Lung analog:     air loss in alveoli -> tumor fills space -> cancer

SLIDES PROCESSED: {len(vsd_df)}

VSD CHANNEL MEANS:
"""

for ch in VSD_CHANNELS:
    col = f'vsd_{ch}_mean'
    if col in vsd_df.columns:
        report += f'  {ch:<25}: {vsd_df[col].mean():.4f} +/- {vsd_df[col].std():.4f}\n'

report += f"""
DERIVED METRICS:
  composition_entropy      : {vsd_df['vsd_composition_entropy_mean'].mean():.4f}
  air_tumor_ratio          : {vsd_df['air_tumor_ratio'].mean():.4f}
  stroma_invasion_score    : {vsd_df['stroma_invasion_score'].mean():.4f}

PIPELINE PERFORMANCE:
  Avg tiles/slide          : {vsd_df['n_tiles'].mean():.0f}
  Avg reduction            : {vsd_df['reduction_pct'].mean()*100:.1f}%
  Avg time/slide           : {vsd_df['time_s'].mean():.1f}s

OUTPUT FILES:
  {DATA_DIR}/hne_vsd_results.csv
  {FIG_DIR}/vsd_channel_distributions.png
  {FIG_DIR}/vsd_air_vs_stroma.png
  {FIG_DIR}/vsd_entropy_survival.png
"""

report_path = os.path.join(OUTPUT_DIR, 'VSD_SUMMARY_REPORT.txt')
with open(report_path, 'w') as f:
    f.write(report)

print(report)
print(f'Report saved: {report_path}')
```
